# The impact of working during the Covid-19 pandemic on health care workers and first responders: mental health, function, and professional retention

**DOI:** 10.1101/2020.12.16.20248325

**Authors:** Rebecca C. Hendrickson, Roisin A. Slevín, Bernard P. Chang, Ellen Sano, Catherine McCall, Murray A. Raskind

## Abstract

**Background:** The COVID-19 pandemic has greatly affected front line health care workers (HCW) and first responders (FR). The infection risk from SARS CoV-2, the high mortality of hospitalized COVID-19 patients and the duration of the pandemic have created sustained and often traumatic stressors affecting HCW and FR.

**Objectives:** To assess the relationship of COVID-19 stressor frequency scores to psychiatric rating scale scores amongst HCW and FR. To determine if psychiatric rating scale scores mediate stressor effects on perceived work function and likelihood of remaining in current occupation.

**Design:** Observational, self-report in a convenience sample.

**Participants:** 118 HCW and FR caring for COVID-19 patients in the United States.

**Main Measure:** COVID-19 related stressor frequencies were assessed using a 17-item questionnaire. Psychiatric symptoms were assessed with the PTSD Checklist 5 (PCL5), the Patient Health Questionnaire 9 (for depression) (PHQ9), the Insomnia Severity Index (ISI), and the General Anxiety Disorder 7 (GAD7).

**Key Results:** Stressor frequency scores correlated significantly with PCL5 scores (R=.57, p<1e-8), PHQ scores (R=.35, P<.001), ISI scores (R=.38, p<1e-4), and GAD7 scores (R=.39, p<.001), likelihood of staying in current occupation (R=-.39,p<1e-4), and trouble doing usual work (R=.33,p<.001). 51% of HCW and 44% of FR indicated decreased likelihood of staying in their current occupation. PCL5 scores substantially mediated the association between stress frequency scores and work function impairment.

**Conclusions:** These results direct attention to recognizing potentially treatable psychiatric symptoms, particularly those of PTSD, in HCW and FR experiencing COVID-19 related stressors. They also suggest that mitigating COVID-19 related stressors when possible, such as by providing adequate personal protective equipment, can improve HCW and FR mental health, work function and retention in the health care work force.

**Strengths and Limitations of this Study:**

- Detailed assessments of participants’ exposure to covid-19 related occupational stressors, current psychiatric symptoms, and self-reported occupational functioning and likelihood of remaining in their current field (functional outcomes).
- Assessment of the dose-response relationship between exposure to covid-19 related occupational stressors and current psychiatric symptoms and functional outcomes.
- Mediation analysis quantifying the potential for current psychiatric symptoms to mediate the relationship between exposure to covid-19 related occupational stressors and functional outcomes.
- Limitations: convenience sample, limited numbers of first responder participants

## Introduction

The impact of prolonged physical and emotional stress on health care workers (HCW) and first responders (FR) working during the Covid-19 pandemic has attracted significant attention in the media and popular press^1,2^. These discussions have chronicled the impact of pandemic-related occupational stressors on HCW/FR as individuals, and have also brought up the potential significance for the healthcare system more broadly. These concerns are consistent with previous research that has found the physical and emotional stressors among HCW/FR can be associated with significant personal distress^3–8^ as well as decreased professional longevity and poorer patient care^9,10^. There remain significant knowledge gaps, however, in understanding the likely impact of working during the conditions experienced during the Covid-19 pandemic on both the short and long term mental health of HCW/FR, and on professional functioning and retention.

Significant attention has been paid to the direct effects of working during a pandemic on the mental health of HCW. HCW and FR working during a pandemic are often exposed to multiple types of acute and sustained stressors, including fear for their own safety and that of their colleagues and family, repeated exposure to death and suffering, extended separations from family, and prolonged periods of exhaustion and vigilance. Depending on the context, they may also experience forms of moral injury^11^ related to their perception of systemic or community support for their work, and the extent to which their contributions feel effective.

High rates of psychiatric symptoms in HCW working during the Covid-19 pandemic, including symptoms of depression, anxiety, insomnia, and PTSD, are well documented^3–8^. Less information is available regarding the specificity of these symptoms for those with specific exposure to pandemic-related workload or risks, although the available evidence suggests that exposure to pandemic stressors such as quarantine procedures, isolation from social support systems, stigmatization, and risk of infection are associated with a higher incidence of psychiatric symptoms^6,7,12,13^.

Far less is known about the potential for increased workplace attrition and impaired workplace functioning due to COVID-19 related occupational stressors. An unpublished but publicly released survey of clinicians carried out by the Larry A Green center and the Primary Care Collaboration reported that 19% of survey respondents reported that clinicians in their practice had retired early because of COVID-19 or were planning on it^14^, and a study of neurosurgeons reported 20% of those surveyed experienced burnout attributed to COVID-19, and were less likely to pursue their career again if given the choice^15^. A study of nurses in South Korea that used a validated questionnaire to assess nurses’ intention to remain in their current career found that those who had worked with COVID-19 patients or in a COVID-19 division reported numerically lower intention to remain in their field than those who had not, but the results were not statistically significant^16^. Little is known, however about the impact on HCW more broadly, which aspects of pandemic-related occupational stress exposure or associated psychiatric symptoms are associated with expected professional retention, or whether they are associated with self-reported functional impairment.

Finally, first responders, such as police service members, firefighters, and emergency medical technicians (EMTs) are exposed to many of the same pandemic-related stressors as hospital-or clinic-based HCW, in even less controlled environments, and with less information about the risk of any given encounter. However, comparatively little information is available regarding the impact of pandemic-related stressors on this population. A review of medical leave in New York City firefighters and emergency medical services (EMS) workers during the early part of the Covid-19 pandemic found increased use of medical leave, leading to decreased workforce availability^17^. A survey of HCW and EMS workers in Italy during the Covid-19 pandemic found that while both groups experienced high levels of distress, FR experienced greater anger and regret, increased intrusiveness related to trauma, and a decreased perception of self-efficacy^18^. However, little or no information is available addressing the exposure to pandemic-related workload in first responders, nor the long-term mental health effects of frontline exposure in these positions.

The existing literature leaves a number of important gaps that impede the ability of the health care system to most effectively address the impact of pandemic-related stressors on its workforce. First, the impact of pandemic related occupational stressors on self-reported functional impairment and likelihood of leaving one’s field, both directly and via the emergence of different types of psychiatric symptoms, is poorly understood. Additional information about these relationships would allow improved targeting of interventions designed to protect HCW and the health care system. Additionally, the focus on traditionally-defined HCW, to the exclusion of FR groups, means little information is available about the needs of or risks to this population.

In this study, we address these existing gaps in the literature in several ways. We assess rate of depression, anxiety, insomnia and PTSD symptoms in both traditionally-defined HCW, as well as in FR (including police, firefighters and EMTs) working during the Covid-19 pandemic. Using an assessment of exposure to occupational stress designed specifically for health care workers and first responders working during the Covid-19 pandemic, we assess the relationship of these symptoms to pandemic-related occupational stressors. Finally, we assess self-reported likelihood of leaving one’s current field and functional impairment when working, and the relationship of these outcomes to both occupational exposure to Covid-19 related stressors, and associated psychiatric symptoms.

## Methods

### 1. Participants

Participants were a convenience sample recruited through targeted outreach and paid advertising on Facebook. Survey responses were completed between September 15 and Nov 24, 2020. Participants were asked to self-attest that they were between the ages of 18-75; that they were a physician (MD or DO), nurse or physician assistant (RN, LPN, ARNP, PA), or first responder (EMT, firefighter, police service member); and that they were or expected to be providing professional services affected by the Covid-19 pandemic. The study was approved by the VA Puget Sound Health Care System Human Subjects Committee. Prior to enrollment, all participants were provided with an electronic-format information statement that detailed the purpose, risks, benefits, and alternatives to participation.

### 2. Procedures

All assessments were self-report, entered electronically using a FEDRamp-adherent implementation of the commercial survey system Qualtrics. Surveys were implemented so as to be responded to on either a computer or any smartphone with a web browser installed. To encourage broad and representative participation despite the sensitive nature of the topic, participants were not required to provide their legal name and were able to skip questions, although a pop-up window notified them of unmarked items and asked if they wished to complete the missing items or continue. Participants were required to provide an email at which they could be contacted for follow up surveys (data not included in this publication). Participants were not compensated for participation.

### 3. Measures

Demographic information collected included age and occupation. Symptoms of depression, anxiety, insomnia, and PTSD were assessed using standard measures: the Patient Health Questionnaire 9-item (PHQ9)^19^, the Generalized Anxiety Disorder 7-item (GAD7)^20^, the Insomnia Severity Index (ISI)^21^, and the PTSD Checklist for DSM-5 (PCL5)^22^, respectively.

Self-reported functional impairment was assessed using the two work-related items from the PROMIS Short Form v2.0 Ability to Participate in Social Roles and Activities 8a measure^23^, modified to focus on occupational work: “I have trouble doing all of my usual work” and “I have trouble doing all of the work that is really important to me”, with answers on a 5-point Likert scale. Likelihood of leaving one’s current profession was assessed with two items: “How likely do you think it is that you will still be working in your current field in 5-10 years?”, with answer choices “not at all likely”, “a little likely”, “moderately likely”, and “highly likely”; and, “How have your experiences providing care during the Covid-19 pandemic affected your interest, willingness, or ability to continue working in your current field?” with answer choices “significantly decreased”, “somewhat decreased”, “no change”, “somewhat increased”, and “significantly increased”.

Exposure to Covid-19 related professional stressors was assessed using a Covid-19 occupational exposure assessment (CEA) survey designed for this study by a cooperative team of physicians working in New York City hospitals and emergency rooms in March and April 2020 and researchers (see Supplemental Table 1). This instrument asks a series of 4 yes/no questions about personal experiences of and loss due to covid-19 infection (scored as 0 or 1), followed by 13 questions addressing the frequency of caring for individuals with Covid-19, perceiving increased risk to self or family due to occupational exposure to Covid-19, experiences of patient suffering related to the impacts of Covid-19, inadequate support or protection related to Covid-19, and feeling unable to provide effective or adequate care due to Covid-19. The 13 frequency questions were asked regarding either the past 2 weeks, if that represented the period of most intense exposure, or retrospectively regarding the period of greatest exposure, with responses “Not at all”, “Several days”, “More than half the days”, and “Nearly every day” (scored as 0-3). The scores were summed to provide a total Covid-19 Exposure Index (CEI).

**Table 1.** Covid-19 occupational exposure assessment. See appendix A for full item content. Responses are indicated as the percent responding positively (the 4 binary response items) or the mean±SD (the 13 Likert scale items). HCW=Health care workers, FR=First responders.

|  | All (N=104) | HCW (N=78) | FR (N=23) |
| --- | --- | --- | --- |
|  | Percent Positive | Percent Positive | Percent Positive |
| Had covid | 31.2 | 27.2 | 44.0 |
| Family member covid | 19.3 | 13.6 | 36.0 |
| Close death from covid | 11.9 | 13.6 | 8.0 |
| At increased risk | 30.5 | 26.9 | 41.7 |
|  | Rating (mean±SD) | Rating (mean±SD) | Rating (mean±SD) |
| Caring mild Sx | 1.60±1.60 | 1.73±1.73 | 1.04±1.04 |
| Covid without PPE | 2.12±2.12 | 2.09±2.09 | 2.20±2.20 |
| Futile care | 1.50±1.50 | 1.63±1.63 | 1.08±1.08 |
| Unable quality for all | 1.16±1.16 | 1.21±1.21 | 0.92±0.92 |
| Risk contracting covid | 1.19±1.19 | 1.27±1.27 | 0.88±0.88 |
| Family increased risk | 1.49±1.49 | 1.67±1.67 | 0.80±0.80 |
| Unsupported by workplace | 1.55±1.55 | 1.70±1.70 | 1.08±1.08 |
| Family separation | 0.97±0.97 | 1.04±1.04 | 0.64±0.64 |
| Unnecessary risks | 0.95±0.95 | 1.06±1.06 | 0.48±0.48 |
| Caring critically ill | 0.79±0.79 | 0.93±0.93 | 0.40±0.40 |
| Longer hours | 1.12±1.12 | 1.26±1.26 | 0.64±0.64 |
| Covid deaths | 1.35±1.35 | 1.57±1.57 | 0.60±0.60 |
| Death in isolation | 2.19±2.19 | 2.16±2.16 | 2.32±2.32 |
| Total Score | 18.0±9.6 | 18.9±9.5 | 14.1±9.6 |

### 4. Data analysis

Data analyzed using R and RStudio. Correlation coefficients were implemented using the *stat_cor* function from the *ggpubr* package. Pearson’s R was used for variables representing scale totals, while Spearman’s correlation coefficients were used when one or more of the variables represented a single ordinal item. Mediation analyses were implemented using the *mediation* package, with statistical significance assessed via bootstrapping using 10,000 simulations. Figures used *ggplot2*. Responses with missing data were excluded from analyses using those data, but included where data was present for all analyzed measures. For all analyses presented, CEI score was based on period of highest exposure.

## Results

### 1. Participants

A baseline questionnaire was completed by 118 participants, with 98% completing all insomnia questions, 86% completing all PTSD questions, 90% completing the exposure questionnaire, and 97% completing the occupation and demographics questions. 87 participants were health care workers (42 physicians, 42 RN or LPN, 4 PA or ARNP; some respondents were in more than one category), while 27 participants were first responders (12 police, 6 firefighters, 11 EMT). Age ranged from 24 to 61 years, with a mean ± standard deviation of 41±9.1.

### 2. Exposure to Covid-19 related occupational stressors

104 participants completed the covid-19 occupational exposure assessment (Table 1). Of these, 31% reported they had been ill with known or likely Covid-19 (27% of HCW and 44% of FR), 19% reported a close family member had been ill with known or likely Covid-19 (36% of FR), 12% reported the death of a family member or close colleague to Covid-19, and 30% indicated they had medical condition which placed them at increased risk from Covid-19. Mean responses to the 13 Likert scale items for the two weeks of highest exposure (total range per item: 0-3) ranged from 0.79±1.05 to 2.19±0.95.

### 3. Impact on psychiatric symptoms

Symptoms of psychiatric disorders that are common after a traumatic stress were quantified using the PCL5 (for PTSD), PHQ9 (for depression), GAD7 (for anxiety) and ISI (for insomnia). 26% of participants had a total PCL5 score of 31 or higher to indicate PTSD symptoms likely to benefit from treatment; 60% had a total PHQ9 score above the standard threshold for at least mild depression, 28% had an ISI score above the threshold for at least moderate insomnia, and 67% had a GAD7 score above the threshold for at least mild anxiety (Table 2).

**Table 2.** Psychiatric symptoms. Mean+/SD of measure totals: PCL5=PTSD Checklist for DSM5, PHQ9=Patient Health Questionnaire 9-item (depression symptoms), ISI=Insomnia Severity Index, GAD7=Generalized Anxiety Disorder 7-item. “% pos” indications percentage over threshold for clinically relevant symptom burden, using total score cutoffs of: 31 for PCL5, 5 for PHQ9, 15 for ISI, 5 for GAD7.

|  | PCL5 Total | % pos | PHQ9 Total | % pos | ISI Total | % pos | GAD7 Total | % pos |
| --- | --- | --- | --- | --- | --- | --- | --- | --- |
| All | 22.31±17.22 | 26.47% | 7.21±5.52 | 60.44% | 11.13±5.66 | 27.59% | 7.77±5.72 | 67.37% |
| HCW | 23.08±16.16 | 26.67% | 7.13±5.04 | 64.18% | 10.59±5.5 | 23.53% | 8.1±5.94 | 68.57% |
| FR | 20±21.24 | 29.17% | 7.45±7.13 | 45.45% | 12.48±5.34 | 40.74% | 6.5±5.3 | 59.09% |

To assess the relationship of psychiatric symptoms to covid-19 related stressors, the relationship of each total symptom score to the CEI was quantified using a Pearson’s correlation coefficient (Figures 1 and S1). Among health care workers specifically, the PCL5 was the most strongly associated with CEI, with an R of .56 (p=4e-7); PHQ9, ISI and GAD7 were related to CEI with R values of .29, .38 and .31 respectively (all p<.05). For first responders (N=23), PCL5, PHQ9, ISI and GAD7 were all strongly associated with CEI (R values of .62, .56, .61 and .58 respectively, all p<.01).

**Figure 1.**
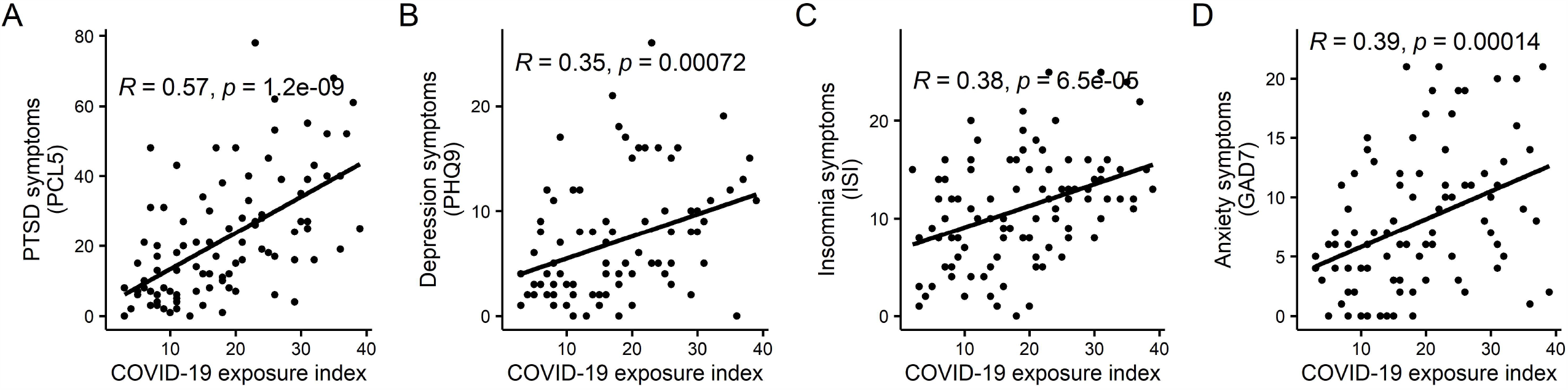
COVID-19 related occupational exposure is strongly related to increased burden of psychiatric symptoms. Covid-19 exposure index (CEI) represents the sum of all responses to the Covid-19 occupational exposure assessment questionnaire. R values represent the Pearson’s coefficient relating the CEI to PTSD symptoms (A), depression symptoms (B), insomnia symptoms (C) and anxiety symptoms (D).

### 4. Impact on self-reported work functioning and likelihood of continuing in current field

Impact on work functioning and likelihood of continuing in one’s current field were captured with four questions (Figure 2). Responses from HCW and FR were similar, with a majority of each group responding they were either “highly likely” or “moderately likely” to be working in their current field in 5-10 years, while approximately a quarter reported this was not at all likely. When asked how their experiences providing care during the Covid-19 pandemic had affected their interest, willingness or ability to continue working in their current field, 11% of HCW and 4% FR reported it had somewhat or significantly increased, 38% of HCW and 52% of FR reported no change, and 51% of HCW and 44% of FR reported it had decreased (20% and 12%, respectively, reporting significantly decreased). 53% of HCW and 44% of FR reported they were sometimes, usually or always unable to do all of their usual work (for HCW, 10% usually and 5% always). Similar or higher numbers reported being unable to “all the work that is really important to me”.

**Figure 2.**
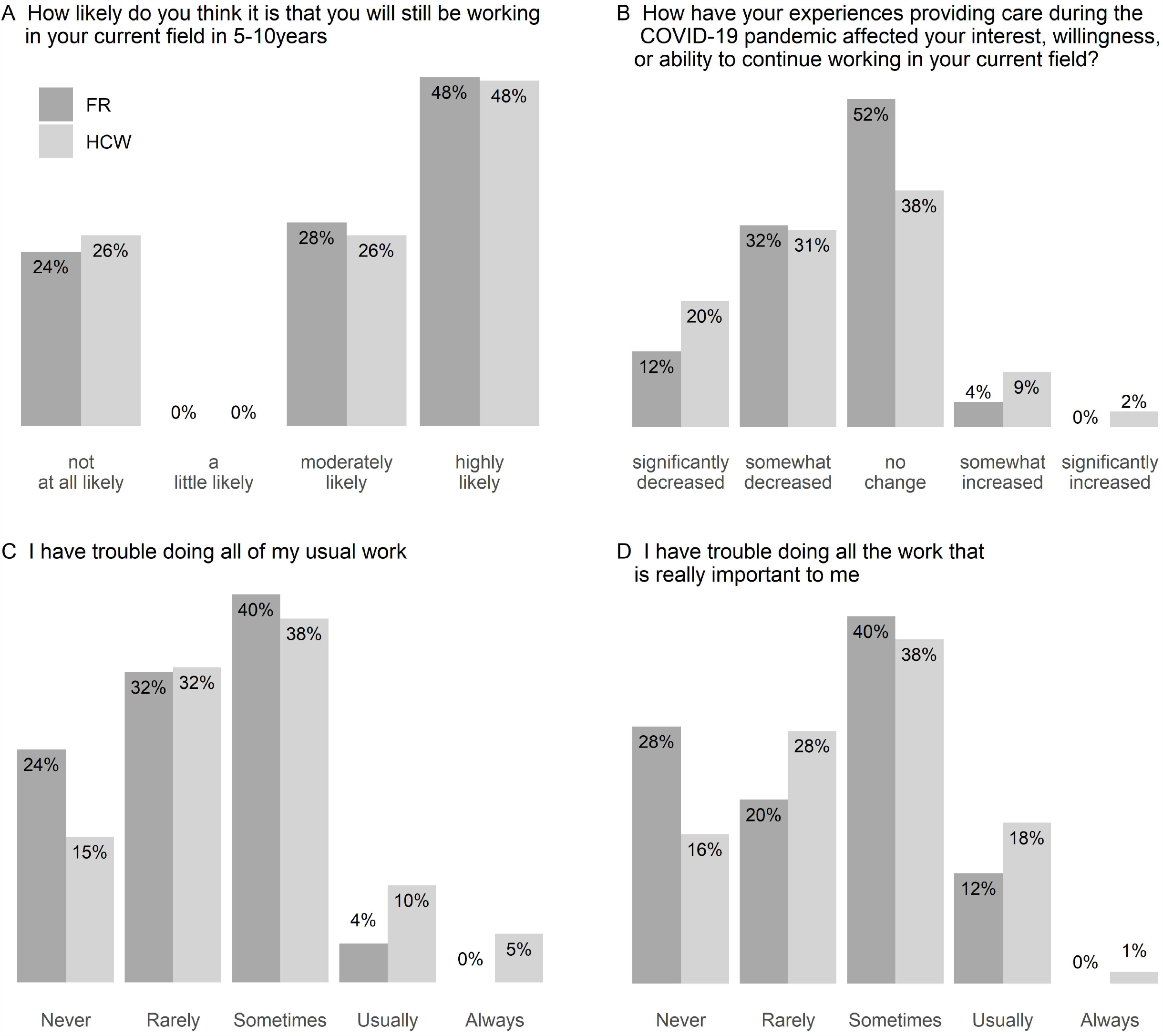
Distributions of responses to self-reported in field retention and functioning. Responses to questions about participants’ expectations regarding continuing in their current field (A,B) and current function (C,D) for health care workers (HCW, n=80) and first responders (FR, n=25).

The relationship of functional impact questions to CEI was assessed via Spearman’s correlation (Figures 3 and S2). Across all participants, likelihood of remaining in one’s current field was rated lower for those with higher CEI, while frequency of being unable to complete one’s usual work was higher. When analyzed for the groups individually, these relationships were statistically significant for the HCW group, but less consistent and not statistically significant for the FR group alone.

**Figure 3.**
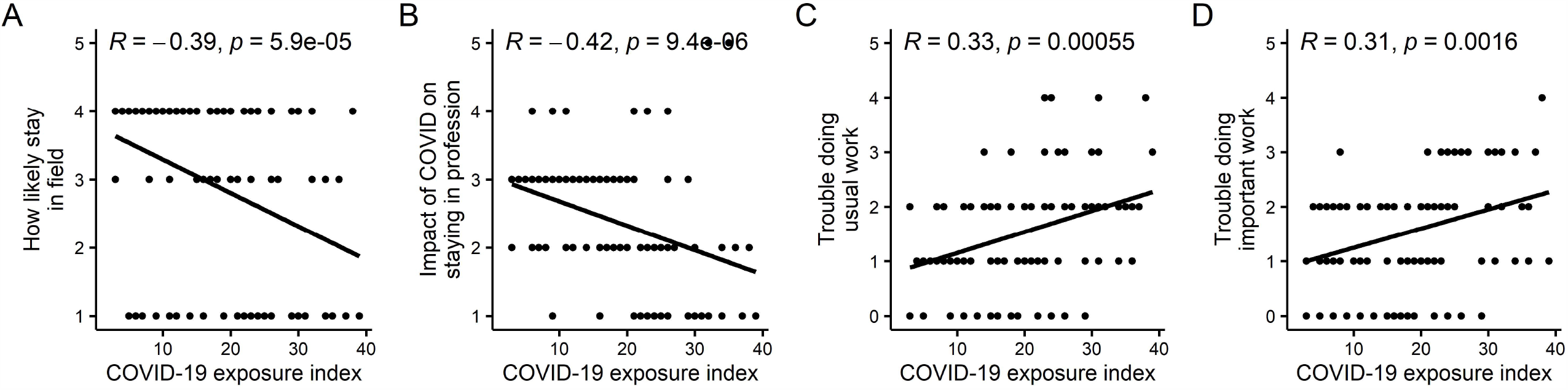
Covid-19 related occupational exposure is strongly related to self-reported likelihood of leaving one’s current field and self-reported impairments in work functioning. Covid-19 exposure index (CEI) represents the sum of all responses to the Covid-19 occupational exposure assessment questionnaire. R represents Spearman’s correlation coefficient.

The role of psychiatric symptom clusters in mediating self-reported functional impairment or likelihood of leaving one’s current field was also assessed (Figure 4); for this analysis, the results were corrected using a Bonferroni correction for n=16 multiple comparisons. A small but statistically significant mediation by insomnia symptoms was identified in relating CEI to likelihood of staying in field. A stronger and more consistent effect was seen relating psychiatric symptoms to inability to complete usual work: PTSD symptoms (PCL5 total) was found to be a highly significant (p<.001 after correction) mediator for both questions regarding work, accounting for the majority of the effect, with insomnia symptoms (ISI total) showing smaller but statistically significant mediation for both work-related items (p<.05 and p<.001, after correction, for ability to complete usual work and important work, respectively).

**Figure 4.**
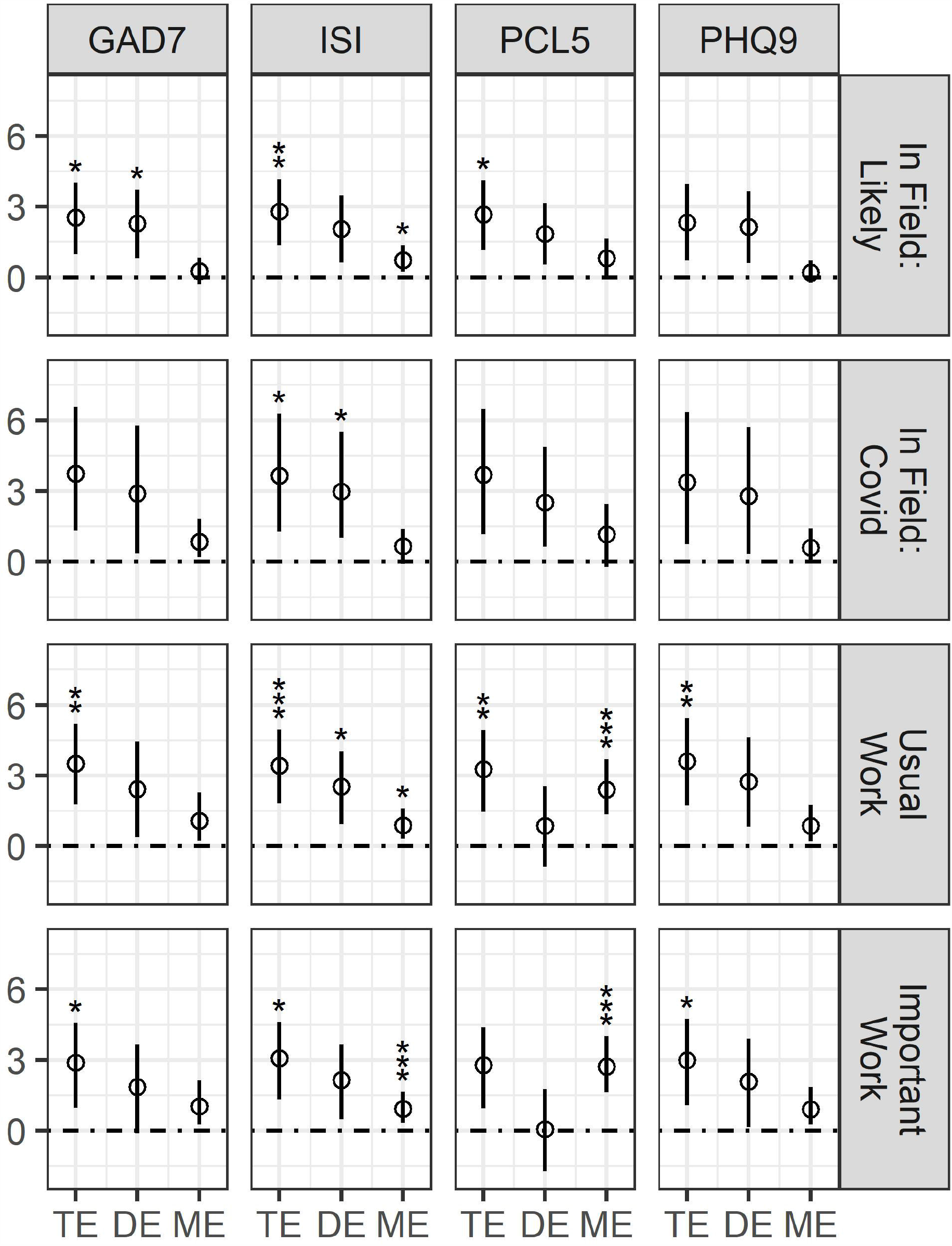
Role of psychiatric symptoms in mediating functional impact of Covid-19 related occupational exposure. Panels show the total effect (TE) and direct effect (DE) of Covid-19 related occupational exposure (CEI) on the four questions addressing likelihood of staying in field and work functioning, as well as the portion of the TE mediated by each type of psychiatric symptoms (ME). Error bars indicate 95% CI, *p<.05, **p<.01, ***p<.001 all adjusted for multiple comparison (N=16).

## Discussion

Consistent with previous work, health care workers reported high levels of psychiatric symptoms after working during the covid-19 pandemic^3–8^. Here, we build on this work by providing an assessment of the relationship of these symptoms to covid-19 related occupational stressors, their presence in FR, and the consequences of these experiences for occupational retention and functioning.

We found that these symptoms were strongly related to a quantitative measure of covid-19 related occupational stress, suggesting they are unlikely to represent baseline rates of psychiatric symptoms in this population during this time period, and are instead more specifically related to experiences working during the pandemic. Although the total number of first responder participants remains low at this time, the available responses showed very similar degrees of distress and were also strongly related to the index of covid-19 related occupational stress. These results suggest that the experiences of first responders should be considered in future work addressing the impact of pandemic related occupational stressors on our society in general and health and safety related systems in particular.

A substantial proportion of both groups reported their likelihood of staying in their current field had been somewhat or significantly decreased by their experiences working during the Covid-19 pandemic, and that they at least sometimes have trouble completing all of their usual or important work. In both cases, these findings were strongly related to the index of occupational exposure. Surprisingly, however, the relationship of participants’ self-reported likelihood of leaving their current field to covid-19 related occupational exposure was only minimally mediated by psychiatric symptoms. This may mean that the impact of occupational stress due to working during the covid-19 pandemic on retention in health care and related fields will not be effectively addressed by either preventing or treating psychiatric symptoms related to stressful experiences, but will instead require what are likely to be system-level changes in order to alter individuals’ experiences working in these fields – such as the availability of PPE, workplace support, sense of efficacy in caring for patients during such periods, or total exposure.

The finding of increased likelihood of leaving one’s current field in HCW with increased COVID-19 related occupational stress is consistent with and builds upon findings from previous pandemics. Almost a tenth of HCW surveyed in a SARS-affected hospital felt reluctance to work or had considered resigning^24^. Another assessment of workplace behavior in healthcare workers who had provided hospital care during a SARS outbreak found decreased total work hours, increased numbers of missed work shifts, and decreased patient contact^25^.

In contrast to the findings related to expected retention in one’s current field, participants’ self-reported work-related functioning was strongly related to PTSD symptom burden, with an additional significant relationship to insomnia symptoms. This suggests that prioritizing the prevention and/or treatment of insomnia and particularly PTSD symptoms may be important for protecting workplace functioning during such periods of prolonged occupational stress.

The prominence of sleep-related symptoms and their relationship to both current function and likelihood of staying in field is also consistent with prior research. Insomnia symptoms were identified in a study of medical staff caring for patients with COVID-19 in China as related to social support with anxiety, stress and self-efficacy as mediating variables^12^. Sleep disruption or insomnia during or immediately following a traumatic stressor has been identified as a significant risk factor for the development of chronic PTSD symptoms^26,27^, and it is hypothesized that this disruption may play a significant and causal role in the development of PTSD over time^28^.

Studies of the impact of working during previous pandemics also underscores the need for longitudinal follow up of the affected workforce. After the SARS outbreak, meta-analysis found that nearly a quarter of HCW met criteria for clinically significant posttraumatic stress disorder (PTSD) during the acute phase of the outbreak^29^, with half that percentage still reporting PTSS over 12 months after the outbreak subsided. HCW in Toronto experienced more long-term adverse psychiatric outcomes following a 2003 SARS outbreak than their counterparts in hospitals not treating SARS patients^25^. Further work following the trajectory of the outcome measure in the current work over time will be important, including to identify forms of pandemic-related occupational stressors that most acutely increase long term risk, and to identify individuals most likely to benefit from directed intervention or longer term support.

The current work has significant limitations. The number of first responders included is low, and replication in a larger sampling will be required. Additionally, the use of targeted outreach and paid advertising targeting regions with high rates of Covid-19 cases for recruitment, rather than systematic sampling of a defined population, means that the absolute rates of exposure to pandemic-related occupational stressors and of psychiatric symptoms reported in the population sampled should not be presumed to reflect rates in all health care workers, nationally or internationally.

In addition, the assessment of covid-19 related occupational stress used in this study has not been validated. It is expected that future work, using a larger total number of respondents, will be able to provide psychometric validation of the exposure questionnaire. Particularly important will be the completion of a factor analysis, and an assessment of which aspects of pandemic-related occupational stressors are most closely related to both psychiatric symptoms and self-reported functional impairment and likelihood of leaving one’s current field. Such additional information could guide more targeted interventions to change health care workers’ and first responders’ experiences during such period of increased stress.

Finally, although insomnia and particularly PTSD symptoms were found to mediate the relationship between occupational exposure and self-reported functional impairment, suggesting that prevention and/or treatment of insomnia and PTSD symptoms in particular may be able to improve self-reported functioning, this hypothesis will need to be tested in a prospective trial.

The impact of the covid-19 pandemic on our health care and first responder systems has been nearly unprecedented in its breadth and duration. As our work and others have shown, many HCW/FR have experienced significant psychiatric distress during this period. Here, we have shown a close connection between the intensity of covid-19 related occupational stressors and not just psychiatric symptoms experienced but also self-reported functional impairment and increased likelihood of leaving one’s current field. We found that PTSD symptoms in particular strongly mediated the relationship between these stressors and functional impairment, suggesting that the assessment and treatment of PTSD symptoms may be particularly important for workers exposed to covid-19 related occupational stressors, but that psychiatric symptoms played a smaller role in mediating the relationship between these stressors and decreased expectations of remaining in one’s current field. Finally, we found that these effects impacted the health care and first responder work force broadly, including groups that have previously received less attention than medical professionals. Future work focused on clarifying these relationships may point to targeted interventions to best support our frontline workers during this pandemic and during future events.

### Funding

This work was supported by a Research & Development Seed Grant from the Department of Veterans Affairs (VA) Puget Sound Health Care System (RCH), VA Clinical Sciences Research and Development Service Career Development Award IK2CX001774 (RCH); and the VA Northwest Network MIRECC (RCH, MAR). BPC is supported by funding from the National Institutes of Health (R01 HL 141811, HL R01 HL 146911).

## Data Availability

De-identified data may be requested by contacting the corresponding author.

## Acknowledgements

The authors would like to thank Dr. Ronald Thomas for brief statistical consultation, and the many participants who generously shared their time and experiences with us. We would also like to acknowledge the significant number of staff members who contributed to this study often beyond their usual work roles, including Carolyn L Fort, Joseph D Clark, Kim Hart, Adam T McPartlin, Emma C Onstad-Hawes, Soleil S Groh, and Andrew Nicholls.

## Contributions of authors

RCH designed the study, with input from all authors. RCH supervised the implementation of the study, with RAS playing a significant role in all implementation steps. RCH carried out the analysis of results, with consultation by Dr. Ronald Thomas. RCH wrote the initial draft of the manuscript, with contributions from RAS and MAR. All authors provided input into the interpretation of the analysis and the drafting of the manuscript.

## Supplementary figure legends

**Supplementary figure 1.**
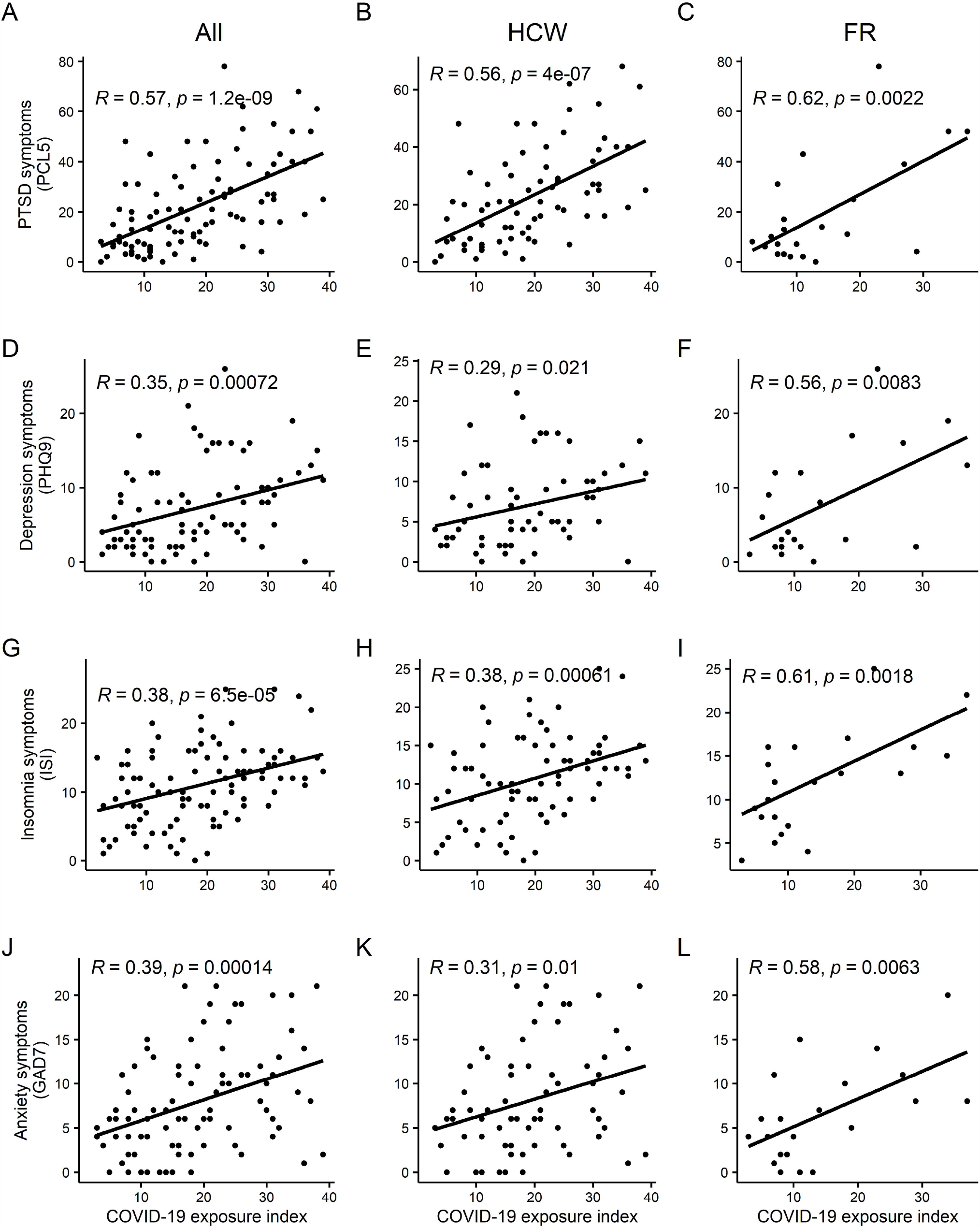
COVID-19 related occupational exposure is strongly related to increased burden of psychiatric symptoms across both HCW and FR, whether the groups are analyzed together or independently. Covid-19 exposure index (CEI) represents the sum of all responses to the Covid-19 occupational exposure assessment questionnaire. R values represent the Pearson’s coefficient relating the CEI to PTSD symptoms (A-C), depression symptoms (D-F), insomnia symptoms (G-I) and anxiety symptoms (J-L).

**Supplementary figure 2.**
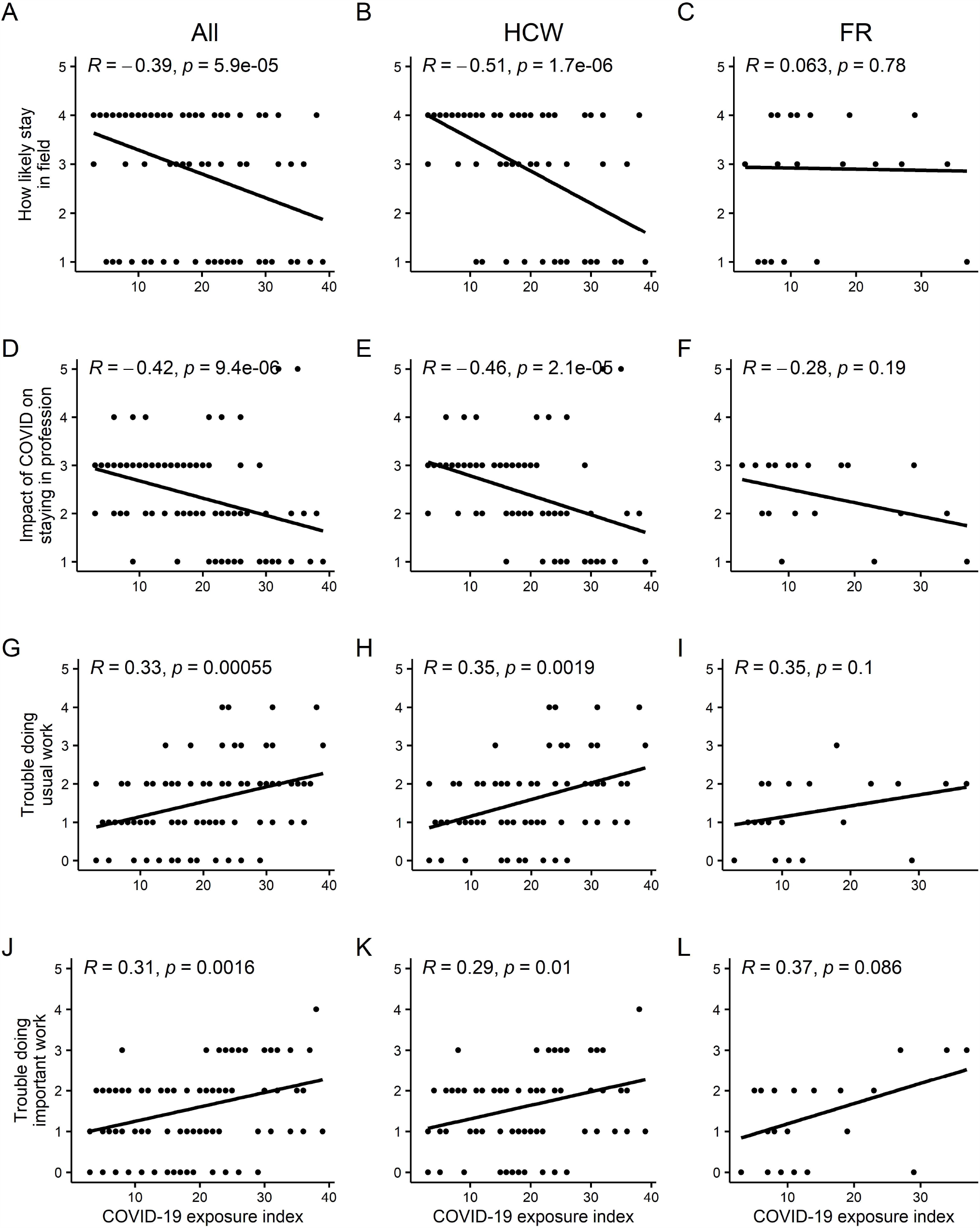
Covid-19 related occupational exposure is strongly related to self-reported likelihood of leaving one’s current field and self-reported impairments in work functioning in HCW and when HCW and FR are analyzed together, but many of these relationships are not maintained in the FR group when analyzed alone. Covid-19 exposure index (CEI) represents the sum of all responses to the Covid-19 occupational exposure assessment questionnaire. R represents Spearman’s correlation coefficient.

## Appendix A. COVID-19 Occupational Exposure Assessment (CEA) for health care workers and first responders

1. *Binary questions, asked without reference to a specific time period:* **At any point, have you experienced the following?** *(Yes, No)*

1. Had a known or strongly suspected infection with COVID-19
2. Had a family member that you live with or see regularly had a known or strongly suspected infection with COVID-19
3. Experienced the death of family member or close colleague from known or suspected COVID-19
4. **Do you have medical conditions (not age alone) that put you at higher risk from COVID-19?** *(Yes, No)*
2. *Likert scale questions, asked with reference to the 2 week period with the greatest degree of distress, disruption or risk related to COVID-19, or the past 2 weeks if this represents the most intense period of exposure:* **In the following questions, we will ask about your work-related experiences with individuals known or strongly suspected to have COVID-19**. Please apply these questions to your own work experiences as a health care worker or first responder. For example, if a question asks about “caring for patients” and you are a first responder, please include any experiences you have responding to calls, even if you would not normally use the word “patient”. **How often did you experience the following:** *(0=Not at all, Several Days, More than half the days, 3=Nearly every day)*

5. Caring for someone with mild symptoms related to COVID-19
6. Caring for someone critically ill with COVID-19
7. Working longer hours than usual in order to provide assistance or care to individuals with COVID-19
8. Witnessing or responding to a death related to COVID-19, or losing a patient you had been caring for to COVID-19
9. Caring for patients who have died without family physically present due to COVID-19 precautions
10. Caring for patients with known or suspected COVID-19 when you were not able to utilize appropriate PPE for the situation
11. Feeling your care for patients with COVID-19 has been futile or unhelpful
12. Feeling unable to provide high quality care to all patients
13. Feeling at increased risk for contracting COVID-19 due to your work
14. Feeling your family was at increased risk for contracting COVID-19 due to your work
15. Feeling unsupported by your workplace
16. Maintaining separation from a member of your household in order to reduce the risk to those you live with
17. Being expected to do things professionally that unnecessarily increase your risk of exposure to COVID-19?

